# Linear age-course association between non-alcoholic fatty liver disease, components of metabolic syndrome, and mortality

**DOI:** 10.1101/2023.08.01.23293528

**Authors:** Xinyu Zhao, Shuohua Chen, Xiao mo Wang, Zhenyu Wang, Yuanyuan Sun, Chenlu Yang, Mengyi Zheng, Yanhong Wang, Wei Liao, Shouling Wu, Li Wang

**Affiliations:** Department of Epidemiology and Biostatistics, Institute of Basic Medical Sciences Chinese Academy of Medical Sciences; School of Basic Medicine Peking Union Medical College, Beijing, China; Department of Clinical Epidemiology and EBM, Beijing Friendship Hospital, Capital Medical University; National Clinical Research Center for Digestive Diseases, Beijing, China; Department of Cardiology, Kailuan General Hospital, Tangshan, China; Cardiovascular Center, Beijing Tongren Hospital, Capital Medical University, Beijing, China

**Author notes:** Corresponding author: Li Wang Department of Epidemiology and Biostatistics, Institute of Basic Medical Sciences Chinese Academy of Medical Sciences; School of Basic Medicine Peking Union Medical College, 5 Dong Dan San Tiao, Beijing 100005, China. E-mail address Shouling Wu Department of Cardiology, Kailuan General Hospital, Tangshan 063000, China. **Abbreviations:** non-alcoholic fatty liver disease (NAFLD); metabolic syndrome (MetS); cardiovascular disease (CVD); elevated blood pressure (BP); systolic blood pressure (SBP); diastolic blood pressure (DBP); elevated fasting blood glucose (FBG); triglyceride (TG); high- density lipoprotein (HDL).

**Keywords:** Non-alcoholic fatty liver disease, metabolic syndrome, mortality, attributable population fraction, cohort.

## Abstract

**Background:** The study aimed to explore the relationship between non-alcoholic fatty liver disease (NAFLD) and mortality by age and metabolic syndrome (MetS) components.

**Method:** We enrolled 104,173 participants in the Kailuan cohort from June 2006 through October 2013. Cox regression models were used to evaluate the hazard ratio (HR) and 95% confidence interval (CI). Population-attributable fractions (PAFs) of metabolic components were also calculated.

**Results:** Compared with non-NAFLD, the excess risk of all-cause and cardiovascular disease (CVD)-related mortality in NAFLD increased with the number of MetS components but decreased with age. The highest all-cause and CVD-related death risks were observed in NAFLD aged 18-39 with 4-5 MetS (HR=2.81, 95% CI:1.55-5.08) and those aged 40-49 years with 3 MetS (HR=2.25, 95% CI:1.55-5.08), respectively. However, there was no significant extra risk of liver-related death in NAFLD patients of any age. For PAF, 28.5% of all-cause and 43.4% of CVD-related mortality among NAFLD patients was preventable by controlling MetS components < 2, with the highest PAF in those aged 50-69 and 18-49, respectively. Elevated blood pressure (PAF of 24.7% for all-cause; 38.6% for CVD-related), elevated fasting glucose (PAF of 13.4% all-cause; 9.2% CVD-related), and elevated triglycerides (PAF of 3.9% all-cause; 14.0% CVD-related) were the essential mortality contributors in NAFLD participants.

**Conclusion:** The excess risk of all-cause and CVD-related mortality in NAFLD patients decreases with age. A substantial proportion of risks could be averted if NAFLD patients are controlled under two MetS components, especially managing blood pressure, fasting glucose, and triglycerides for the young.

## 1. Introduction

Non-alcoholic fatty liver disease (NAFLD) is the most prevalent chronic liver disease worldwide, with a global prevalence of 30%^1^. NAFLD may increase the risk of cardiovascular disease, chronic kidney disease, and some specific cancers^2, 3^. However, its impact on mortality is still controversial due to the difference in the study population’s characteristics and the diagnosis method of NAFLD^4–8^. Most NAFLD-related mortality studies were based on western people or limited by highly selected patient populations^4, 5, 9, 10^.

The participants with NAFLD are often accompanied by components of metabolic syndrome (MetS) . Studies have shown that the risk of all-cause mortality gradually increased with the number of components of MetS^11^. Also, the early onset of the individual component of MetS, such as obesity, diabetes, and hypertension, has been reported to have a higher risk of death than those with late onset ^12–14^. Furthermore, our cohort study in a Northern Chinese population has shown a modified effect of age on the association between NAFLD and mortality^15^. All the above information suggests that age may modify the effect of NAFLD on all-cause mortality. Additionally, it is unclear what extent of deaths can be reduced by controlling MetS components of NAFLD patients to normal levels.

Using the Kailuan cohort in China, we aimed to: (1) evaluate the association between the excess risk of mortality among patients with NAFLD versus non-NAFLD participants by the number of MetS components; (2) quantify the proportions of deaths in the NAFLD and non-NAFLD that can be reduced by controlling different MetS components to normal levels.

## 2. Methods

### 2.1 Study population

Starting from 2006 and being followed up every two years, the Kailuan cohort is an ongoing longitudinal prospective cohort based on the Kailuan community in Tangshan, China. In this study, we enrolled 159,018 participants who entered the cohort between 2006-2013. We excluded 54,804 participants due to the following: (1) having liver cirrhosis at baseline (n=170); (2) excessive alcohol consumption (alcohol intake ≥30 g/day for men and ≥20 g/day for women) or with alcohol intake missing (n=42524); (3) hepatitis B surface antigen (HBsAg) positive or missing (n=4931); (4) missing on the fatty liver by B ultrasound (n=6864); (5) death within one year from the baseline (n=315); or (6) three or more missing values of MetS components (n=41). Finally, 104,173 participants were included (Supplementary Figure 1).

The Ethics Committees of the Kailuan General Hospital and the Institute of Basic Medical Sciences Chinese Academy of Medical Sciences approved this study. We obtained informed consent from all participants.

### 2.2 Definition of NAFLD, MetS, and its metabolism components

Using abdominal ultrasonography (PHILIPS HD-15), fatty liver was diagnosed by experienced radiologists blinded to clinical presentation and laboratory results. Participants with fatty liver were defined as NAFLD because we excluded individuals with excessive alcohol consumption or other causes of chronic liver disease. We utilized the diagnostic criteria proposed by the International Diabetes Federation (2009) ^16^ to define MetS by the presence of ≥3 of the following components: abdominal obesity, elevated blood pressure (BP), fasting blood glucose (FBG) serum triglyceride (TG), and high-density lipoprotein (HDL) (detailed in Supplementary text).

### 2.3 Follow-up and outcome assessment

We followed all the participants to the date of death, or 12-31-2019, which came first. Death certificates were obtained annually from the Kailuan social security system and provincial vital statistics offices. The cause of death listed on the death certificate was classified following the 10^th^ version of the International Classification of Diseases (ICD-10). We defined the cause of death due to cardiovascular disease (CVD) with codes of ICD-10 I00-I69 ^17^ and liver-related mortality with ICD-10 of C22, B15-B19, I85, K70- K76 ^18^. Death causes were collected before 31 December 2016, so the follow-up ended at the date of death or 12-31-2016 when analyzing the risk of CVD-related and liver-related death.

### 2.4 Potential confounders

We conducted questionnaire surveys to collect demographic characteristics (age, sex, and education level), lifestyle factors (smoking, drinking, and physical activity), medical history, and medications of all the participants. Height, weight, waist, and blood pressure were measured by trained field workers. Laboratory tests, including TG, HDL, alanine aminotransferase (ALT), FBG, and HBsAg were assessed in the central lab.

### 2.5 Statistical methods

Variables were presented as mean ± standard deviation or number (%) and compared using analysis of variance or Chi-square test, respectively. The age- and sex- standardized prevalence of MetS component for NAFLD and non-NAFLD populations were estimated using the Chinses 2010 census population as the reference ^19^. We conducted the Kaplan-Meier to calculate the mortality rate. Compared to the non-NAFLD, the Cox proportional hazards model with age as the timescale was used to estimate the HRs and 95% confidence intervals (CIs) of NAFLD with the different numbers of MetS components after adjusting for age, sex, education, smoking, physical activity, salt intake, and ALT level. We divided the participants into six groups (18-39, 40-49, 50- 59, 60-69, 70-79, and ≥80 years) because we found an age-dependent hazard ratio (HR) between NAFLD and death risk (Supplementary Figure 2).We also conducted sensitivity analyses to verify the results: (1) using the original complete dataset, (2) excluding females, (3) excluding drinkers, (4) defining NAFLD as the participants diagnosed with moderate or severe fatty liver for the low sensitivity and specificity of ultrasound in the diagnosis of mild fatty liver ^20, 21^, and (5) excluding participants with cancers or cardiovascular disease (myocardial infarction, stroke, subarachnoid hemorrhage, cerebral infarction, cerebral hemorrhage or heart failure) at baseline.

Furthermore, we assessed the adjusted overall and age-specific population-attributable fraction (PAF) of deaths that contributed to MetS components in the NAFLD and compared them with that in the non-NAFLD using a method for cohort study ^22^. We used multiple imputations by chained equations to impute one complete dataset with five interactions, which showed equal distribution before and after imputation (Supplementary Table 1).

Analyses were implemented with SAS version 9.4 (SAS Institute Inc. Cary, NC), R version 3.5.2 (https://www.r-project.org/), or Stata 15.0.

## 3. Results

### 3.1 Baseline characteristics and the prevalence of metabolic components

Among 104,173 participants with a mean age of 49.2±14.0 years old, NAFLD was identified in 32,384 (31.1%) participants, 74.9% of whom combined with >1 MetS component, significantly higher than in the non-NAFLD (35.7%). In the NAFLD group, the average age and the proportion of people with low education levels, high-intensity physical exercise, salt preference, and high ALT gradually increased with the increasing number of MetS components (Table 1).

**Table 1.** Baseline characteristics of subjects with and without NAFLD.

| Variable | Non-NAFLD | NAFLD |  |  |  | *p*-value |
| --- | --- | --- | --- | --- | --- | --- |
|  |  | G<sub>0-1</sub> | G<sub>2</sub> | G<sub>3</sub> | G<sub>4-5</sub> |  |
| N | 71789 | 8115 | 10229 | 9260 | 4780 |  |
| Age, years | 49.1±14.6 | 48.5±12.9 | 51.6±12.7 | 53.4±12.1 | 54.3±11.5 | <0.001 |
| Male | 52865(73.6) | 6403(78.9) | 8172(79.9) | 7211(77.9) | 3485(72.9) | <0.001 |
| Education |  |  |  |  |  | <0.001 |
| Primary school or below | 6355(8.9) | 541(6.7) | 897(8.8) | 958(10.3) | 555(11.6) |  |
| Junior high school | 47265(65.8) | 5437(67.0) | 7129(69.7) | 6509(70.3) | 3341(69.9) |  |
| Senior high school | 10750(15.0) | 1243(15.3) | 1412(13.8) | 1214(13.1) | 589(12.3) |  |
| College and above | 7419(10.3) | 894(11.0) | 791(7.7) | 579(6.3) | 295(6.2) |  |
| Physical activity level* |  |  |  |  |  | 0.003 |
| Inactive | 8765(12.2) | 1002(12.3) | 1202(11.8) | 1044(11.3) | 534(11.2) |  |
| Moderate | 53227(74.1) | 6127(75.5) | 7623(74.5) | 6843(73.9) | 3397(71.1) |  |
| Frequently | 9797(13.6) | 986(12.2) | 1404(13.7) | 1373(14.8) | 849(17.8) |  |
| Smoking status |  |  |  |  |  | <0.001 |
| Never | 52061(72.5) | 5692(70.1) | 7150(69.9) | 6484(70.0) | 3367(70.4) |  |
| Past | 2923(4.1) | 341(4.2) | 584(5.7) | 552(6.0) | 270(5.6) |  |
| Current | 16805(23.4) | 2082(25.7) | 2495(24.4) | 2224(24.0) | 1143(23.9) |  |
| Salt intake |  |  |  |  |  | <0.001 |
| Low (<6g/d) | 7391(10.3) | 790(9.7) | 982(9.6) | 905(9.8) | 484(10.1) |  |
| Medium (6-11g/d) | 58454(81.4) | 6627(81.7) | 8200(80.2) | 7429(80.2) | 3731(78.1) |  |
| High (>11g/d) | 5944(8.3) | 698(8.6) | 1047(10.2) | 926(10.0) | 565(11.8) |  |
| ALT (U/L) |  |  |  |  |  | <0.001 |
| <40 | 69126(96.3) | 7279(89.7) | 8917(87.2) | 7898(85.3) | 3943(82.5) |  |
| 40-79 | 2246(3.1) | 713(8.8) | 1099(10.7) | 1138(12.3) | 697(14.6) |  |
| 80- | 417(0.6) | 123(1.5) | 213(2.1) | 224(2.4) | 140(2.9) |  |

Data are presented as mean (standard deviation) or number (percentage).
\*We defined physical activity as inactive, moderately active, and active regarding the frequency of physical activity (≥20 min/time) during leisure time with responses of never, sometimes, and ≥ four times/week, respectively.
Abbreviations: NAFLD: Non-alcoholic fatty liver disease; G<sub>0-1</sub>, G<sub>2</sub>, G<sub>3</sub>, G<sub>4-5</sub>: with 0-1, 2, 3, and 4-5 metabolic syndrome components, respectively; ALT, aminotransferase.

The standardized prevalence of each MetS component in the NAFLD group was significantly higher than that in the non-NAFLD group (Table 2). The highest difference in prevalence between NAFLD and non-NAFLD was abdominal obesity (62.7% *vs.* 27.3%), then the elevated TG (47.5% *vs.* 18.4%) and elevated BP (50.9% *vs.* 30.7%).

**Table 2.** Standardized prevalence of metabolic syndrome components stratified by NAFLD. non-NAFLD NAFLD.

| MetS components | non-NAFLD |  | NAFLD |  | *P* |
| --- | --- | --- | --- | --- | --- |
|  | n/N | Standardized prevalence (%) | n/N | Standardized prevalence (%) |  |
| Total population |  |  |  |  |  |
| Elevated FBG | 16935/71789 | 20.0 | 13166/32384 | 36.9 | <0.001 |
| Elevated BP | 27109/71789 | 30.7 | 19147/32384 | 50.9 | <0.001 |
| Abdominal Obesity | 21063/71789 | 27.3 | 20889/32384 | 62.7 | <0.001 |
| Elevated TG | 14245/71789 | 18.4 | 16184/32384 | 47.5 | <0.001 |
| Reduced HDL | 7821/71789 | 15.1 | 4738/32384 | 21.8 | <0.001 |
| G<sub>0-1</sub> | 46167/71789 | 68.2 | 8129/32384 | 29.0 | <0.001 |
| G<sub>2</sub> | 16746/71789 | 21.0 | 10197/32384 | 30.5 | <0.001 |
| G<sub>3</sub> | 7054/71789 | 8.5 | 9285/32384 | 26.5 | <0.001 |
| G<sub>4-5</sub> | 1822/71789 | 2.3 | 4773/32384 | 14.0 | <0.001 |

Standardized prevalence: based on the direct standardization method using the Chinses 2010 census population as the reference;
Elevated FBG: administration of anti-diabetes drugs or fasting blood glucose $\geq 5.6$ mmol/L; elevated BP: administration of anti-hypertensive drugs and(or) systolic blood pressure(SBP) $\geq 140$ mmHg or diastolic blood pressure(DBP) $\geq 90$ mmHg; abdominal obesity: waist circumference $> 90$ cm(male) or $> 85$ cm(female); elevated TG: TG $\geq 1.7$ mmol/L; reduced HDL: HDL $< 1.0$ mmol/L(male) or $< 1.3$ mmol/L(female).
Abbreviations: NAFLD: Non-alcoholic fatty liver disease; MetS: metabolic syndrome; FBG: fasting blood glucose; BP: blood pressure; TG: triglycerides; HDL: high-density lipoprotein cholesterol; G<sub>0-1</sub>, G<sub>2</sub>, G<sub>3</sub>, G<sub>4-5</sub>: with 0-1, 2, 3, and 4-5 metabolic syndrome components, respectively.

### 3.2 Associations between NAFLD with the different numbers of MetS components and CVD-, liver-related, and all-cause mortality

During a median follow-up of 12.7 years, 10442 subjects died. The 10-year all-cause mortality rates (Figure 1A) of the non-NAFLD (7.5%) were significantly lower than the NAFLD with 3 (10.3%) and 4-5 MetS components (11.6%) but higher than the NAFLD with 0-1 MetS component (1.5%), *p* <0.001. Similar phenomena were observed in CVD-related death (Figure 1B, *p* < 0.001). Although existing an increased trend, we did not observe a significant difference in liver-related mortality between NAFLD with the different numbers of MetS components and non-NAFLD (Figure 1C, p=0.314).

**Figure 1.**
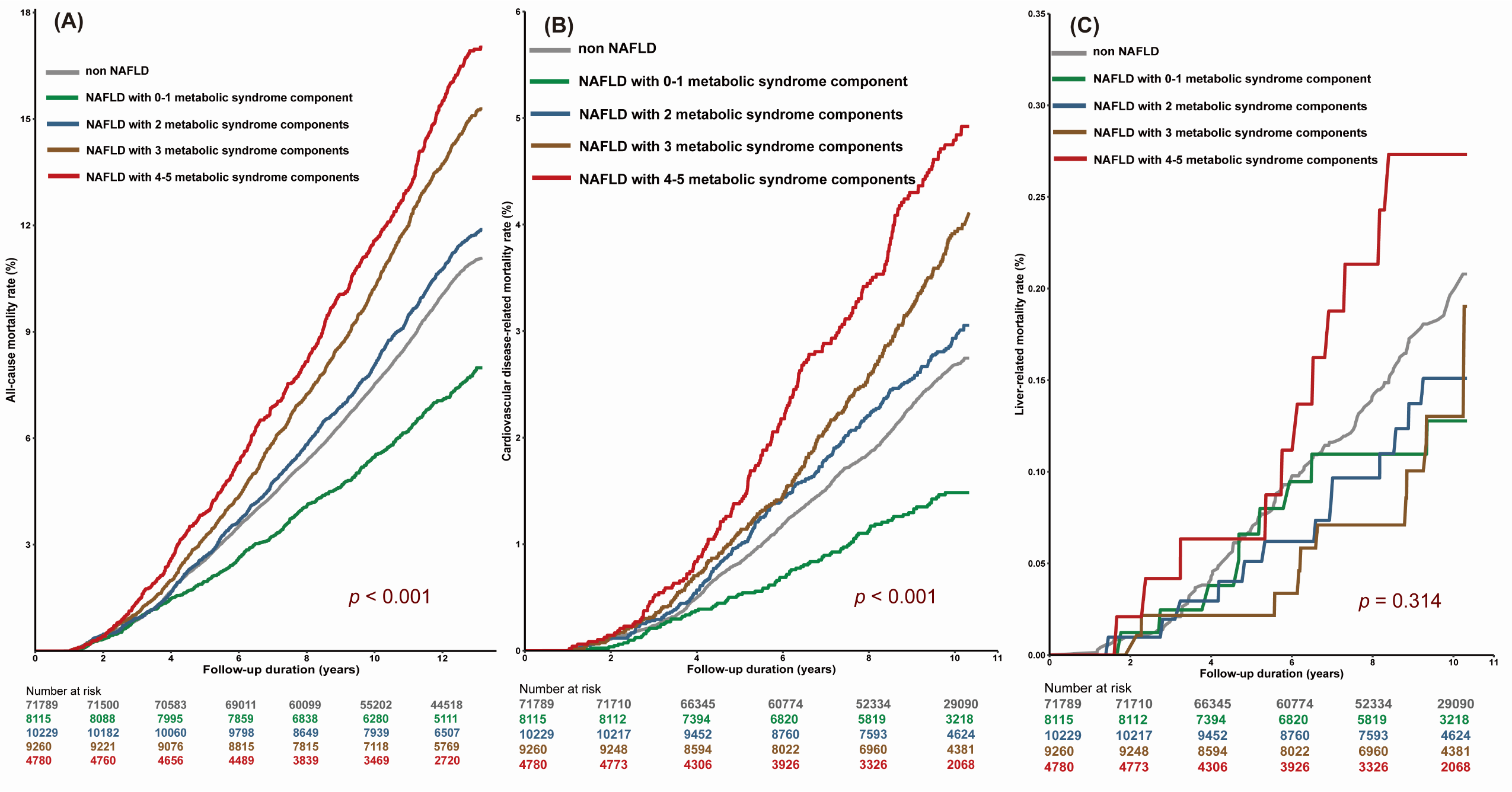
The cumulative all-cause, CVD-, and liver-related mortality rates of participants with different numbers of metabolic syndrome components

### 3.3 Associations between NAFLD with the different numbers of MetS components and CVD-, liver-related, and all-cause mortality by age

Figure 2A showed that the excess risk of all-cause mortality for NAFLD vs. non-NAFLD increased with the number of metabolic component abnormalities but linearly declined with age. In the subjects with 4-5 MetS components but aged < 40, NAFLD patients had the highest all-cause mortality risk (HR=2.81, 95% CI:1.55-5.08, *p*=0.001).

**Figure 2.**
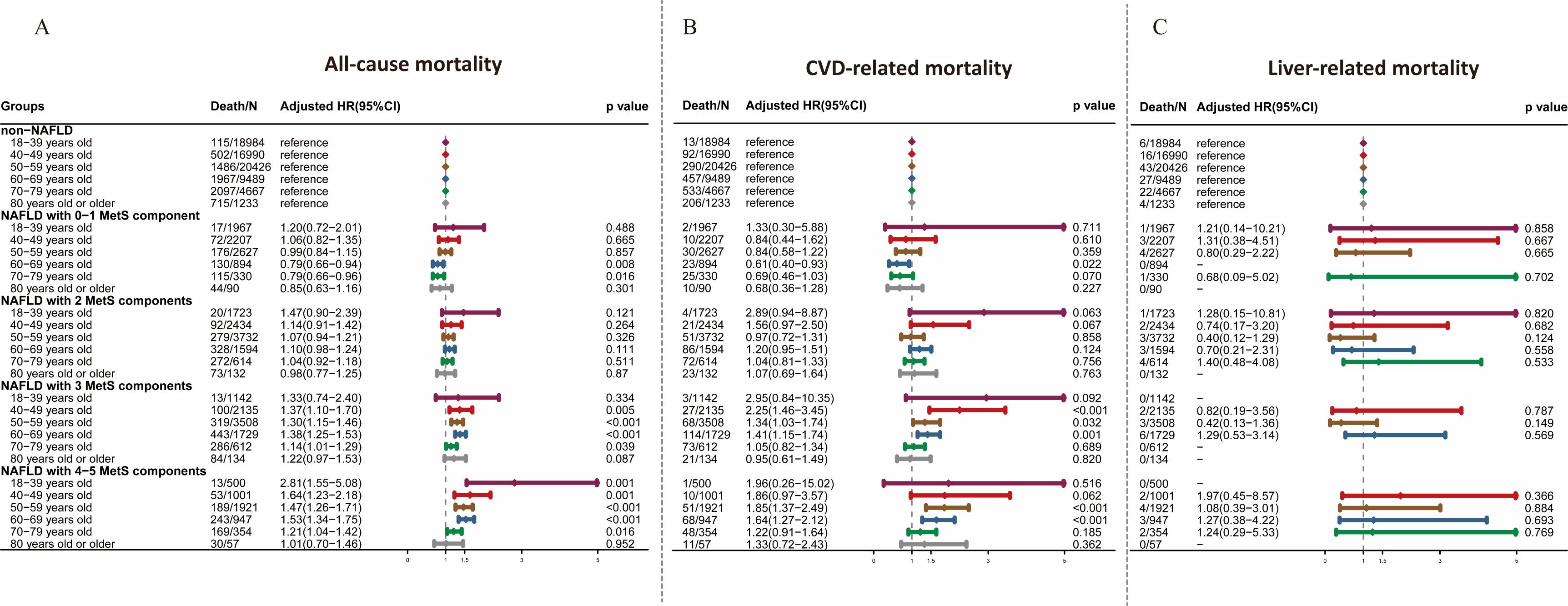
The excess risk of all-cause, CVD-, and liver-related mortality among patients with NAFLD *vs.* non-NAFLD according to the number of metabolic components. Adjusted HRs: Adjusted for sex, education, smoking, physical activity, salt intake, and alanine aminotransferase.

For CVD-related mortality (Figure 2B), higher risks for NAFLD vs. non-NAFLD was only observed in those aged 40-69 and with >2 MetS components. For the NAFLD patients with 3 MetS components, the HR(95% CI) for those aged 30-49, 50-59, and 60-69 was 2.25(1.46-3.45), 1.34(1.03-1.74), and 1.41(1.15-1.74). For NAFLD patients with 4-5 MetS components, the HR(95% CI) at the corresponding age was 1.86 (0.97-3.57), 1.85(1.37-2.49), and 1.64(1.27-2.12). However, we did not observe an excess risk of liver-related death risk in NAFLD patients in any age group (figure 2C). So we did not perform further PAF-related analyses for liver-related death.

The sensitivity analysis showed a consistent result after excluding females, drinkers, individuals with cancers or CVD, NAFLD with mild fatty liver, or using original data (Supplementary Table 2-4).

### 3.4 Association between metabolic components and all-cause and CVD-related death by NAFLD status

Table 3 showed the association between MetS components and all-cause mortality by NAFLD status. Due to the limited outcome events in the NAFLD patients, we regrouped the age category into <50, 50-69, and ≥70 years. Both in the NAFLD and non-NAFLD populations, elevated FBG (HR=1.36, 95% CI:1.27-1.46 in the NAFLD; HR=1.29, 95% CI:1.23-1.36 in the non-NAFLD) and elevated BP (HR=1.49, 95% CI:1.38-1.62 in the NAFLD; HR=1.39, 95% CI:1.32-1.47 in the non-NAFLD) were significantly associated with all-cause death after multivariable adjustments. Elevated TG (HR=1.08, 95% CI:1.01-1.16) was only observed to be a risk factor for death in the NAFLD, and abdominal obesity in the non-NAFLD (HR=1.07, 95% CI:1.01-1.12). However, HRs of elevated BP on death declined with age in both NAFLD and non-NAFLD groups. HR(95%CI) decreased from 1.66(1.32-2.09) for those aged 18-49 to 1.21(1.04-1.40) for those aged ≥70 in the NAFLD and from 1.66 (1.40-1.98) to 1.12(1.03-1.21) in the non-NAFLD correspondingly.

**Table 3.**
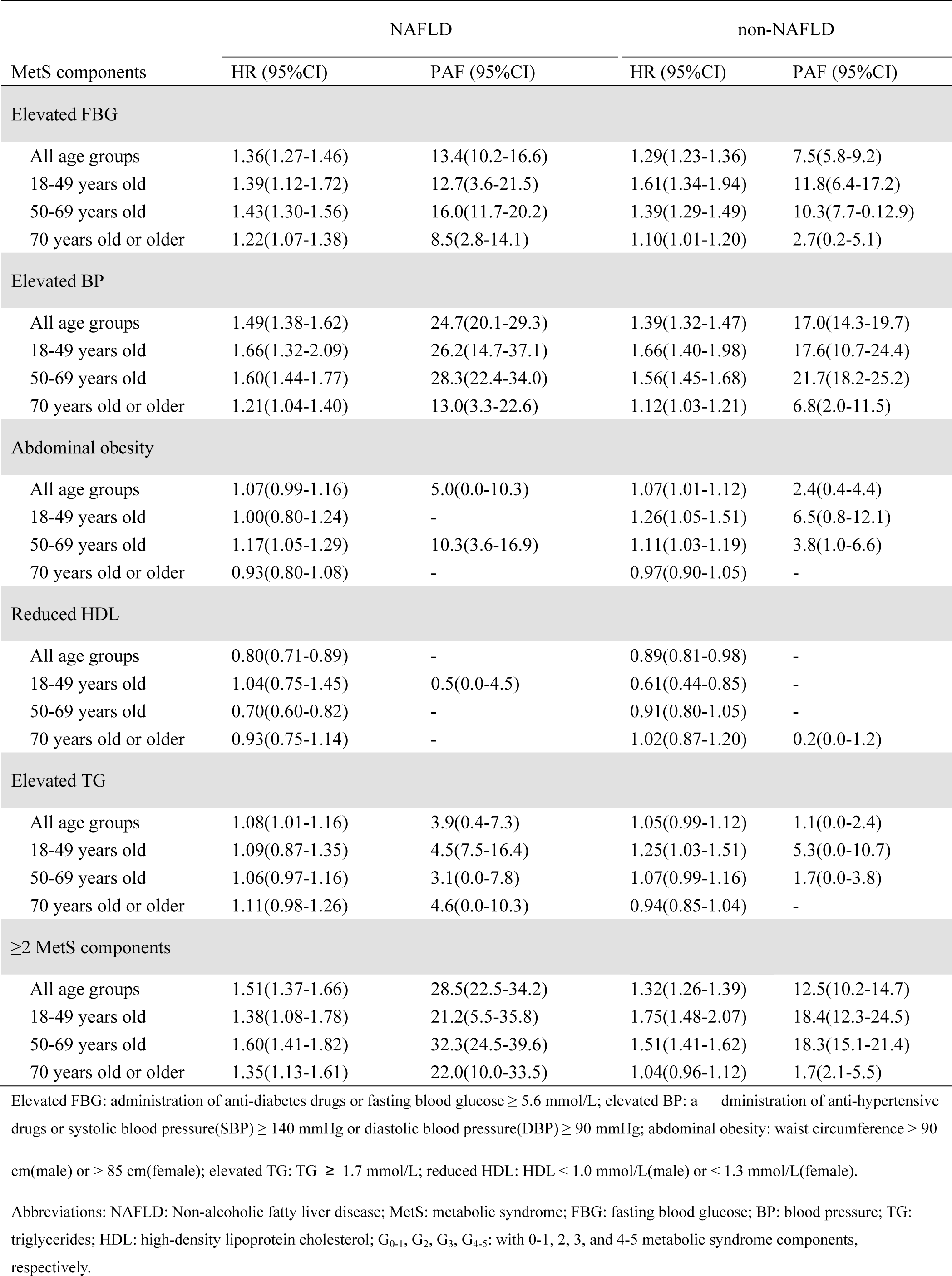
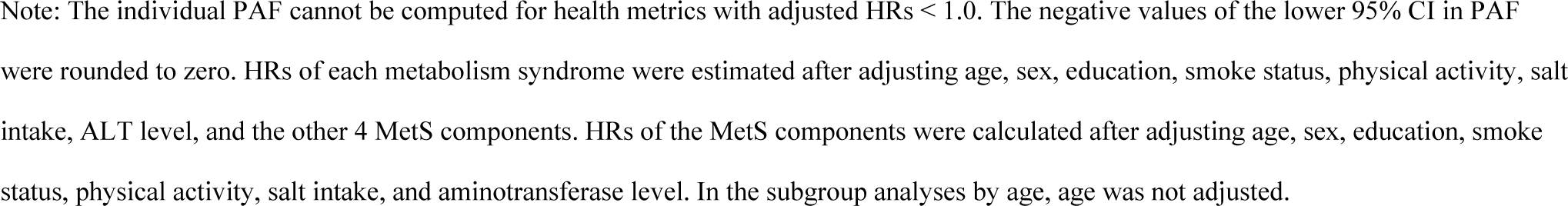
Adjusted hazard ratios and population-attributable fractions (95% CI) of metabolic components to all-cause mortality by NAFLD status.

| MetS components | NAFLD |  | non-NAFLD |  |
| --- | --- | --- | --- | --- |
|  | HR (95%CI) | PAF (95%CI) | HR (95%CI) | PAF (95%CI) |
| Elevated FBG |  |  |  |  |
| All age groups | 1.36(1.27-1.46) | 13.4(10.2-16.6) | 1.29(1.23-1.36) | 7.5(5.8-9.2) |
| 18-49 years old | 1.39(1.12-1.72) | 12.7(3.6-21.5) | 1.61(1.34-1.94) | 11.8(6.4-17.2) |
| 50-69 years old | 1.43(1.30-1.56) | 16.0(11.7-20.2) | 1.39(1.29-1.49) | 10.3(7.7-0.12.9) |
| 70 years old or older | 1.22(1.07-1.38) | 8.5(2.8-14.1) | 1.10(1.01-1.20) | 2.7(0.2-5.1) |
| Elevated BP |  |  |  |  |
| All age groups | 1.49(1.38-1.62) | 24.7(20.1-29.3) | 1.39(1.32-1.47) | 17.0(14.3-19.7) |
| 18-49 years old | 1.66(1.32-2.09) | 26.2(14.7-37.1) | 1.66(1.40-1.98) | 17.6(10.7-24.4) |
| 50-69 years old | 1.60(1.44-1.77) | 28.3(22.4-34.0) | 1.56(1.45-1.68) | 21.7(18.2-25.2) |
| 70 years old or older | 1.21(1.04-1.40) | 13.0(3.3-22.6) | 1.12(1.03-1.21) | 6.8(2.0-11.5) |
| Abdominal obesity |  |  |  |  |
| All age groups | 1.07(0.99-1.16) | 5.0(0.0-10.3) | 1.07(1.01-1.12) | 2.4(0.4-4.4) |
| 18-49 years old | 1.00(0.80-1.24) | - | 1.26(1.05-1.51) | 6.5(0.8-12.1) |
| 50-69 years old | 1.17(1.05-1.29) | 10.3(3.6-16.9) | 1.11(1.03-1.19) | 3.8(1.0-6.6) |
| 70 years old or older | 0.93(0.80-1.08) | - | 0.97(0.90-1.05) | - |
| Reduced HDL |  |  |  |  |
| All age groups | 0.80(0.71-0.89) | - | 0.89(0.81-0.98) | - |
| 18-49 years old | 1.04(0.75-1.45) | 0.5(0.0-4.5) | 0.61(0.44-0.85) | - |
| 50-69 years old | 0.70(0.60-0.82) | - | 0.91(0.80-1.05) | - |
| 70 years old or older | 0.93(0.75-1.14) | - | 1.02(0.87-1.20) | 0.2(0.0-1.2) |
| Elevated TG |  |  |  |  |
| All age groups | 1.08(1.01-1.16) | 3.9(0.4-7.3) | 1.05(0.99-1.12) | 1.1(0.0-2.4) |
| 18-49 years old | 1.09(0.87-1.35) | 4.5(7.5-16.4) | 1.25(1.03-1.51) | 5.3(0.0-10.7) |
| 50-69 years old | 1.06(0.97-1.16) | 3.1(0.0-7.8) | 1.07(0.99-1.16) | 1.7(0.0-3.8) |
| 70 years old or older | 1.11(0.98-1.26) | 4.6(0.0-10.3) | 0.94(0.85-1.04) | - |
| ≥2 MetS components |  |  |  |  |
| All age groups | 1.51(1.37-1.66) | 28.5(22.5-34.2) | 1.32(1.26-1.39) | 12.5(10.2-14.7) |
| 18-49 years old | 1.38(1.08-1.78) | 21.2(5.5-35.8) | 1.75(1.48-2.07) | 18.4(12.3-24.5) |
| 50-69 years old | 1.60(1.41-1.82) | 32.3(24.5-39.6) | 1.51(1.41-1.62) | 18.3(15.1-21.4) |
| 70 years old or older | 1.35(1.13-1.61) | 22.0(10.0-33.5) | 1.04(0.96-1.12) | 1.7(2.1-5.5) |

Elevated FBG: administration of anti-diabetes drugs or fasting blood glucose $\geq 5.6$ mmol/L; elevated BP: a dministration of anti-hypertensive drugs or systolic blood pressure(SBP) $\geq 140$ mmHg or diastolic blood pressure(DBP) $\geq 90$ mmHg; abdominal obesity: waist circumference $> 90$
cm(male) or $> 85$ cm(female); elevated TG: TG $\geq 1.7$ mmol/L; reduced HDL: HDL $< 1.0$ mmol/L(male) or $< 1.3$ mmol/L(female).
Abbreviations: NAFLD: Non-alcoholic fatty liver disease; MetS: metabolic syndrome; FBG: fasting blood glucose; BP: blood pressure; TG: triglycerides; HDL: high-density lipoprotein cholesterol; G<sub>0-1</sub>, G<sub>2</sub>, G<sub>3</sub>, G<sub>4-5</sub>: with 0-1, 2, 3, and 4-5 metabolic syndrome components, respectively.
Note: The individual PAF cannot be computed for health metrics with adjusted HRs $< 1.0$ . The negative values of the lower 95% CI in PAF were rounded to zero. HRs of each metabolism syndrome were estimated after adjusting age, sex, education, smoke status, physical activity, salt intake, ALT level, and the other 4 MetS components. HRs of the MetS components were calculated after adjusting age, sex, education, smoke status, physical activity, salt intake, and aminotransferase level. In the subgroup analyses by age, age was not adjusted.

Regarding CVD-related death (Table 4), the results were similar to all-cause mortality. The risk effect of elevated BP (HR=1.94, 95% CI:1.63-2.30 in the NAFLD; HR=1.65, 95% CI:1.49-1.84 in the non-NAFLD) and evaluated TG ((HR=1.33, 95% CI:1.16-153 in the NAFLD; HR=1.13, 95% CI:1.01-1.27 in the non-NAFLD) on CVD-related death was more prominent than that on the all-cause mortality. And the risk effect of evaluated TG also declined with age. HR(95%CI) decreased from 1.58(0.99-2.55) to 1.24(0.98-1.57) in the NAFLD group and from 1.56(1.02-2.37) to 1.03(0.85-1.24) in the non-NAFLD group.

**Table 4.** Adjusted hazard ratios and population-attributable fractions (95% CI) of metabolic components to CVD-related mortality by NAFLD status.

| MetS components | NAFLD |  | non-NAFLD |  |
| --- | --- | --- | --- | --- |
|  | HR (95%CI) | PAF (95%CI) | HR (95%CI) | PAF (95%CI) |
| **Elevated FBG** |  |  |  |  |
| All age groups | 1.23(1.07-1.41) | 9.2(3.0-15.4) | 1.33(1.20-1.48) | 8.5(5.1-12.0) |
| 18-49 years old | 1.18(0.75-1.85) | 6.4(0.0-24.5) | 2.04(1.36-3.05) | 18.9(5.2-32.0) |
| 50-69 years old | 1.30(1.09-1.56) | 12.2(3.7-20.5) | 1.51(1.31-1.76) | 13.6(8.1-19.0) |
| 70 years old or older | 1.08(0.85-1.37) | 3.4(0.0-14.0) | 1.05(0.89-1.23) | 1.3(0.0-5.8) |
| **Elevated BP** |  |  |  |  |
| All age groups | 1.94(1.63-2.30) | 38.6(29.9-46.8) | 1.65(1.49-1.84) | 25.9(20.5-31.1) |
| 18-49 years old | 2.48(1.46-4.19) | 44.9(20.4-64.1) | 1.73(1.17-2.57) | 19.9(3.3-35.5) |
| 50-69 years old | 2.21(1.76-2.78) | 44.6(33.5-54.4) | 2.25(1.92-2.63) | 38.4(31.2-45.1) |
| 70 years old or older | 1.37(1.03-1.82) | 20.9(2.7-37.9) | 1.14(0.98-1.33) | 8.0(0.0-17.0) |
| **Abdominal obesity** |  |  |  |  |
| All age groups | 1.09(0.93-1.27) | 6.0(0.0-16.7) | 1.08(0.98-1.20) | 3.0(0.0-7.1) |
| 18-49 years old | 0.90(0.57-1.43) | - | 1.44(0.96-2.17) | 11.0(0.0-24.5) |
| 50-69 years old | 1.18(0.97-1.45) | 11.2(0.0-24.3) | 1.11(0.96-1.29) | 4.2(0.0-10.1) |
| 70 years old or older | 1.00(0.75-1.33) | - | 0.99(0.86-1.15) | - |
| **Reduced HDL** |  |  |  |  |
| All age groups | 0.76(0.60-0.97) | - | 0.80(0.65-0.99) | - |
| 18-49 years old | 0.71(0.31-1.64) | - | 0.52(0.23-1.19) | - |
| 50-69 years old | 0.71(0.52-0.96) | - | 0.76(0.56-1.03) | - |
| 70 years old or older | 0.88(0.59-1.33) | - | 0.98(0.72-1.32) | - |
| **Elevated TG** |  |  |  |  |
| All age groups | 1.33(1.16-1.53) | 14.0(7.1-20.8) | 1.13(1.01-1.27) | 2.7(0.0-5.5) |
| 18-49 years old | 1.58(0.99-2.55) | 24.0(0.0-47.4) | 1.56(1.02-2.37) | 11.7(0.0-25.6) |
| 50-69 years old | 1.34(1.12-1.61) | 15.0(5.6-24.3) | 1.14(0.96-1.34) | 3.2(0.0-7.6) |
| 70 years old or older | 1.24(0.98-1.57) | 9.7(0.0-20.4) | 1.03(0.85-1.24) | 0.5(0.0-4.0) |
| **≥2 MetS components** |  |  |  |  |
| All age groups | 1.97(1.60-2.43) | 43.4(32.3-53.3) | 1.43(1.29-1.58) | 16.3(11.7-20.8) |
| 18-49 years old | 2.36(1.27-4.38) | 48.7(17.2-71.2) | 2.30(1.57-3.39) | 28.3(13.6-41.8) |
| 50-69 years old | 2.16(1.62-2.87) | 47.9(33.5-60.1) | 1.78(1.54-2.06) | 25.6(19.1-31.8) |
| 70 years old or older | 1.51(1.06-2.15) | 29.4(0.0-49.5) | 1.05(0.91-1.21) | 2.3(0.0-9.4) |

Elevated FBG: administration of anti-diabetes drugs or fasting blood glucose $\geq 5.6$ mmol/L; elevated BP: administration of anti-hypertensive drugs and(or) systolic blood pressure(SBP) $\geq 140$ mmHg or diastolic blood pressure(DBP) $\geq 90$ mmHg; abdominal obesity: waist circumference $> 90$ cm(male) or $> 85$ cm(female); elevated TG: TG $\geq 1.7$ mmol/L; reduced HDL: HDL $< 1.0$ mmol/L(male) or $< 1.3$ mmol/L(female).
Abbreviations: CVD: cerebrovascular disease; NAFLD: Non-alcoholic fatty liver disease; MetS: metabolic syndrome; FBG: fasting blood glucose; BP: blood pressure; TG: triglycerides; HDL: high-density lipoprotein cholesterol; G<sub>0-1</sub>, G<sub>2</sub>, G<sub>3</sub>, G<sub>4-5</sub>: with 0-1, 2, 3, and 4-5 metabolic syndrome components, respectively.
Note: The individual PAF cannot be computed for health metrics with adjusted HRs $< 1.0$ . The negative values of the lower 95% CI in PAF were rounded to zero. HRs of each metabolism syndrome were estimated after adjusting for age, sex, education, smoke status, physical activity, salt intake, ALT level, and the other 4 MetS components. HRs of MetS components were calculated after adjusting age, sex, education, smoke status, physical activity, salt intake, and aminotransferase level. In the subgroup analyses by age, age was not adjusted.

### 3.5 PAFs of metabolic components to all-cause/CVD-related mortality among NAFLD and non-NAFLD patients

The prevalence of MetS components by age was shown in Supplementary Table 5. The MetS components increased with age, except for the reduced HDL in the NAFLD and non-NAFLD participants.

Table 3 showed the PAFs of MetS components to all-cause death by NAFLD status. For both NAFLD and non-NAFLD participants, the largest adjusted PAF of MetS components was elevated BP (24.7% in the NAFLD and 17.0% in the non-NAFLD), and then the elevated FBG (13.4% in the NAFLD and 7.5% in the non-NAFLD). With age increasing, the PAF of elevated BP (from 26.2% to 13.0% in the NAFLD and from 17.6% to 6.8% in the non-NAFLD) and elevated FBG (from 12.7% to 8.5% in the NAFLD, and from 11.8% to 2.7% in the non-NAFLD) were both declined.

Like all-cause of death, elevated BP had the largest adjusted PAF for CVD-related death in both NAFLD patients (38.6%) and non-NAFLD (25.9%) (Table 4). However, the elevated TG (14.0%) had a slightly higher PAF than the elevated FBG (9.2%) for NAFLD patients. The elevated FBG (8.5%) still had the second-largest PAF for non-NAFLD patients. Similarly, the PAFs of elevated BP (from 44.9% to 20.9% in the NAFLD and from 19.9% to 8.0% in the non-NAFLD) and elevated TG (from 24.0% to 9.7% in the NAFLD, and from 11.7% to 0.5% in the non-NAFLD) dramatically declined with age.

For the multiple MetS components, 28.5% of all-cause deaths and 43.4% of CVD-related deaths could be potentially averted by controlling the subjects with NAFLD to combining less than one MetS component, more than twice in the subjects without NAFLD (all-cause mortality: 12.5%; CVD-related mortality: 16.3%).

## 4. Discussion

Our study found that the standardized prevalence of abdominal obesity, elevated BP, TG, and FBG, and reduced HDL reached 62.7%, 50.9%, 47.5%, 36.9%, and 21.8% among NAFLD patients, respectively. Compared to non-NAFLD, The highest all-cause and CVD-related death risks were observed in NAFLD aged 18-39 with 4-5 MetS (HR=2.81, 95% CI:1.55-5.08) and those aged 40-49 years with 3 MetS (HR=2.25, 95% CI:1.55-5.08). Higher than in non-NAFLD, all-cause of death could be attributable to 24.7% of elevated BP and 13.4% of elevated FBG in the NAFLD population. Similarly, CVD-related death could be attributable to 38.6% of high BP, 9.2% of elevated FBG, and 14.0% of elevated TG.

Our analysis showed that patients with NAFLD aged 18-39 with 4-5 MetS components had the highest excess risk of all-cause mortality compared with non-NAFLD. Notably, we identified a monotonic relationship between the increasing age of NAFLD patients and lowered extra risk of all-cause mortality and CVD-related mortality, which could be supported by the research on the early onset of hypertension ^12^, obesity ^13^, and diabetes ^14^ to the excess deaths or CVD-related mortality.

The present study indicated that 28.5% of all-cause deaths and 43.4% of CVD-related deaths among NAFLD patients could be attributable to presenting ≥2 MetS component, suggesting that reducing MetS components <1 may substantially reduce mortality, consistent with a previous study using the NHANES III data ^23^. Furthermore, our study showed that 24.7% of the all-cause and 38.6% of CVD-related mortality in NAFLD participants were attributable to elevated BP, slightly lower than a study from the NHANES III data (adjusted PAF of 23% of all-cause and 52.8% of CVD-related mortality) ^23^. With a prevalence of elevated BP of 50.9% in our NAFLD population, hypertension is an independent driver of the progression of NAFLD to non-alcoholic steatohepatitis, advanced liver fibrosis, cirrhosis, and hepatocellular carcinoma ^24, 25^. Also, CVD risk has been proven to be increased with more advanced liver disease from a meta-analysis of 36 longitudinal studies, furtherly supporting our results^2^. Our former study found that even in the NAFLD with normal BMI, those with hypertension had a higher risk of all-cause death and CVD ^26^, suggesting the importance of controlling blood pressure for NAFLD participants.

Elevated FBG was found to be attributed to 13.4% of all-cause mortality in NAFLD, higher than that in non-NAFLD (7.5%). The higher prevalence of elevated FBG in the NAFLD (36.9%) than that in non-NAFLD (20.0%) could explain the difference since the similar HR for elevated FBG and all-cause mortality in the NAFLD (HR=1.36) and non-NAFLD (HR=1.29) group. Similar results were observed for CVD-related death (PAF: 13.4% in NAFLD vs. 7.5% in non-NAFLD). Diabetes could promote NAFLD progression to cirrhosis and fibrosis and increase the all-cause mortality risk^27, 28^. Insulin resistance could explain the higher risk of all-cause and CVD-related mortality in the participants with elevated FBG.

Elevated TG was found to be the second-largest contributor to CVD-related death (PAF=14%) in NAFLD patients, higher than that of elevated FBG (9.2%). It can be explained by the significant risk effect (elevated TG: HR=1.33 *vs.* elevated FBG: HR=1.23) and higher prevalence (elevated TG: 47.5% *vs.* elevated FBG: 36.9%) of elevated TG in NAFLD patients. Serum elevated TG has been suggested to be an indicator of atherogenic dyslipidemia. Together with hyperglycemia, oxidative stress and inflammation activation, dysregulated lipid metabolism could create a pro-atherogenic environment attributing to CVD development ^29^. We also found that the PAFs of elevated BP and elevated TG to all-cause or CVD-related mortality in NAFLD patients declined with age, which could be explained by the decreased HRs of elevated BP and TG with age.

This study has some limitations. Firstly, we did not collect hepatitis C infection information, so patients with hepatitis C were not excluded. However, given the relatively low rate of hepatitis C infection in the Chinese population (1%-3%), the estimated association between NAFLD, MetS, and all-cause mortality has little impact ^30^. Secondly, 80% of the participants were male, so the impact of NAFLD and MetS components on all-cause mortality in females and its potential sex differences needs further exploration. Finally, the association between NAFLD and liver-related mortality needs more cases and longer follow-ups to prove.

## 5. Conclusion

For NAFLD patients combined with more than one MetS component, its excess risk of all-cause and CVD-related mortality decreased with age. Substantial proportions of deaths, especially CVD-related deaths, could be averted if NAFLD patients are controlled under two MetS components, especially controlling elevated blood pressure, fasting glucose, and triglycerides.

## Acknowledgement

The authors thank all the members of the Kailuan Study Team for their contribution and the participants who contributed their data.

## Declarations

### Data availability

The data used or analyzed during this study are included in this article and available from the corresponding author upon reasonable request.

